# What does Serum Glycosylated Apolipoprotein J mean in a heart attack?-----Correlation of serum glycosylated apolipoprotein J with acute myocardial infarction

**DOI:** 10.1101/2023.06.19.23291631

**Authors:** Rao Yao, Mengmeng Ren, Haibin Dong, Hua Wang, Wenjuan Jia, Xiaoning Ding, Kaixuan Fu, Anyi Wang, Xuefeng Zhu, Lei Gong, Lin Zhong

## Abstract

**Background:** There are few myocardial damage markers that could be used to diagnose acute myocardial infarction(AMI) or assess its severity, especially glycosylated apolipoprotein J(ApoJ-Glyc) has demonstrated superiority in cardiomyocytes and animal STEMI models in the early stages of myocardial ischemia(MI). We aimed to excavate the potential role of ApoJ-Glyc as a protein marker in the pathogenesis of AMI in humans and its added value in the evolution of the disease.

**Methods and Results:** ELISA was used to determine the serum concentration of ApoJ-Glyc in 163 patients enrolled by the criteria. Statistical analysis could used to discuss the relationship between ApoJ-Glyc and AMI. Compared to control groups, serum ApoJ-Glyc levels decreased by 36% and 37% in early AMI patients and AMI patients, respectively (P<0.0001), showing a higher discriminant value for early diagnosis and diagnosis of AMI [area under the curve (AUC) : 0.871 and 0.886, P< 0.0001]. For the first time, we demonstrated that ApoJ-Glyc was not statistically significant in the comparative difference between NSTEMI and STEMI groups (P> 0.05). Patients with gradually declining ApoJ-Glyc had a higher Grace Risk Scores. Subsequent studies have also demonstrated that more MACCE did occur with a 6-month follow-up(P<0.05).

**Conclusions:** ApoJ-Glyc which serve as an alarm bell for the detection of early ischaemia, may be a new biomarker for AMI. ApoJ-Glyc can assess the severity of myocardial infarction. The continuous decrease of serum ApoJ-Glyc suggests an increase in the risk of post-AMI ischaemia and the onset of unpredictable MACCE.

**Graphic abstract:** 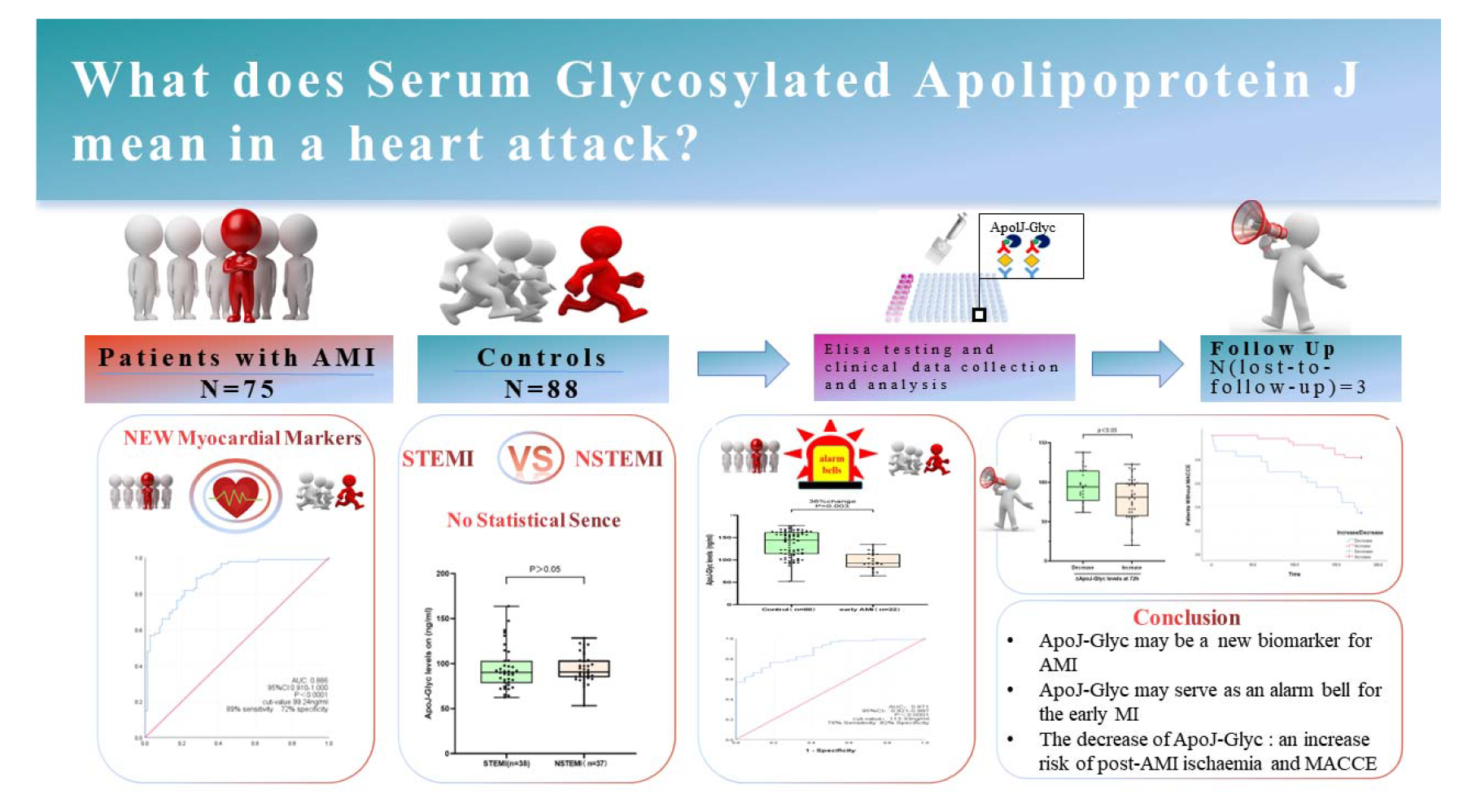

## INTRODUCTION

Cardiovascular disease (CVD) is one of the most common causes of morbidity and mortality in the world^1^.

According to conservative estimates, more than 23.3 million people will die from cardiovascular disease each year by 2030^2^. Internationally, China and India have the highest burden of cardiovascular disease. In China, Cardiovascular disease (CVD) is the leading cause of death and premature death, accounting for 40% of deaths in China^3–5^.

Atherosclerosis, a progressive inflammatory disease characterized by lipid accumulation and cell death within the large-or medium-sized arteries and intimal plaques and cholesterol accumulation in the arterial walls, could lead to ischaemia of the heart, brain, and extremities. In conditions, such as ischemic cardiomyopathy, stable atherosclerotic plaques evolve into unstable fragile plaques, which are prone to rupture and thrombosis, then may eventually lead to acute myocardial infarction^6^. Acute myocardial infarction is one of the most serious complications of cardiovascular disease, which can further cause heart rupture, malignant arrhythmia and severe heart failure, resulting in death of patients^7^. The diagnosis and evaluation of acute coronary syndrome is based on clinical symptoms, electrocardiogram findings, and cardiac-specific troponin (Tn) levels, the most commonly used biomarkers^8, 9^. However, this traditional biomarker has limited effectiveness in the acute phase and its concentration may be too low to detect at the earliest stages^10^.Studies have clearly shown that the release of hs-TnI occurs after completely reversible ischaemia. The delayed release of hs-TnI appears to reflect apoptosis-induced focal cardiomyocyte death and may explain that myocardial hs-TnI is a good marker of irreversible cell damage^11^. However, troponins are elevated in many cardiovascular diseases, such as heart failure, aortic dissection, myocarditis, arrhythmic cardiomyopathy, and atrial fibrillation^12, 13^.As can be seen, important kinetic differences have been observed, cardiac troponin concentrations are insufficient to distinguish between types of myocardial infarction and other causes of myocardial injury or infarction and should not be used alone to guide management decisions^14^.The failure to identify hs-TnI levels and the absence of ischaemia changes on the ECG in patients hospitalized within 1-3 hours of chest pain increase the chance of false negative results, which could lead to extended hospital admissions, needless testing, and a worse prognosis, without demonstrable benefits^15–18^.In this context, a specific ischemic biomarker, combined with hs-TnI, should be able to quickly identify ischemic events in patients with stenosis where Tn levels are undetectable, Tn/ECG results are uncertain, or other indications of low event probability. As a result, there is an ongoing demand for biomarkers to identify patients with AMI in the acute phase. Furthermore, the lack of protein-based biomarkers for risk stratification is also an unmet need that may improve the ability of biomarkers to identify patients at higher risk of adverse outcomes.

Apolipoprotein J (apoJ, also known as clusterin), is a heterodimeric glycoprotein with a molecular weight of about 75-80 kDa, encoded by a single gene located on the short arm of chromosome 8, near the lipoprotein lipase gene locus^19, 20^.As an apolipoprotein, apoJ is found in a subset of dense HDL Particles and exists in two forms: the highly glycosylated secretory form of clusterin (sCLU) and its intracellular nuclear form (nCLU)which is still not well characterized.^21, 22^.Secretory Apol J is considered to be molecular chaperone and its activity depends on the degree of glycosylation^23, 24^. Apolipoprotein J has been found play an important role in lipid transportation and vascular smooth muscle cell (VSMC) differentiation, especially in apoptotic cell death, cell-cycle regulation, tissue remodeling, immune system regulation, and oxidative stress^25^.Clusterin promotes the export of cholesterol and phospholipids in foam cells^26^.The decrease in CLU concentration in the heart muscle tissue is associated with reduction in the degree of its damage and the inhibition of the apoptosis process under conditions of hypoxia or ischaemia^20^.Up to now, most studies on the pathophysiologic function of ApoJ-Glyc in cardiovascular disease have been performed in animal models. Studies have demonstrated that in the porcine STEMI model, serum ApoJ-Glyc levels decreased early after ischemia and were maintained during ischaemia, suggesting that ApoJ-Glyc may be a early and high-sensitive biomarker of acute myocardial ischaemia, which recovered to varying degrees on the third day after myocardial infarction^27^. Currently, the roles and clinical significance of ApoJ-Glyc in humans with AMI are not well established. In this study, we aimed to observe the clinical value of ApoJ-Glyc in patients with AMI.

## MATERIALS AND METHODS

### Study Design

In this study, a total of 163 patients admitted to the emergency department due to typical chest pain were immediately examined by coronary angiography, including 75 patients with acute myocardial infarction underwent coronary stenting (AMI group) and 88 patients without coronary artery stenosis who were included in the control group (Con group). For research purposes and to avoid confounding data, the exclusion criteria are as follows:

1. Patients who have failed PCI;
2. Patients with severe infection and renal insufficiency;
3. Patients with malignant tumors;
4. Patients with combined immune system diseases;
5. patients with severe arrhythmia, heart failure, or cardiac valve problems;
6. Patients with incomplete information, unpredictable accidental death or unable to track and evaluate.

All enrolled patients met the following conditions:

1. Patients admitted to the emergency department due to typical chest pain cannot exclude the possibility of myocardial infarction;
2. Age > 18 years;
3. Complete clinical baseline data;
4. No previous history of myocardial infarction;
5. No previous history of coronary angiography;
6. Each subject signed an informed consent form prior to enrollment;
7. all approved by the Medical Ethics Committee;

The study protocol of this study conforms to the ethical guidelines of the 1975 Declaration of Helsinki.

### ApoJ-Glyc Measurement

Fresh venous blood samples were taken from 163 patients at the time of admission. In 75 patients, additional blood samples were taken 24 and 72 hours after PCI. The samples are processed immediately and stored at −80 °C until ApoJ-Glyc analysis.

### Baseline Parameters

The patients data including medical history and background medication information data were gathered via face-to-face interviews. The same researcher measured the left atrial diameter (LAD), left ventricular end diastolic diameter (LVEDD), LVPW, and left ventricular ejection fraction (LVEF) upon admission using transthoracic echocardiography.

### Evaluation of the TIMI, Gensini and GRACE Score

The TIMI flow classification evaluates the distal flow of the diseased vessel^28^; the Gensini score evaluates the degree of stenosis of the patient’s coronary artery lesion; the GRACE score is a predictive tool for risk stratification rating of patients at admission and discharge from the GRACE study, the Global Registry of Coronary Events, which is recommended by authoritative guidelines both nationally and internationally; the initial assessment should be done within 24 hours of admission and is primarily The first assessment should be completed within 24 hours of admission and is used to predict the risk of death during the patient’s stay in hospital and a review should be performed within 1 week prior to discharge to assess the patient’s prognosis at 6 months after discharge^29^.

### Follow up and survival analysis

Control group: 6 months post-discharge follow-up, no adverse outcomes except for the lost-to-follow-up group (n=3); AMI group: MACCE time recorded up to 6 months after discharge.

### Statistical analysis

We first tested for normality for continuous variables, described by mean ± standard deviation for conformity to normality and t-test (group = 2) or ANOVA (group ≥ 3) for difference type analysis; median and quartiles for non-conformity and value sum test for difference analysis; chi-square test for unordered qualitative data and non-parametric Mann Whitney (=2) or Kruskal-Wallis (>2) tests. Statistically significant variables obtained from the analysis of variance were included in multiple regression analyses (stepwise selection of variables) to assess independent risk factors. To reduce multicollinearity between independent variables, variables were first screened using Lasso regression. Logistic regression analysis was then used to assess the predictive value of ApolJ-Glyc for AMI patients based on lasso regression results. Logistic regression models were assessed using ROC curves.

Bivariate correlations were calculated using the spearman correlation coefficient, and variables that were statistically significant in previous analyses were correlated with ApolJ-Glyc to screen for variables that evaluated the severity of myocardial infarction for analysis of variance and linear regression analysis. For prognostic information Kaplan-Meier curves were used for survival analysis.

LASSO regression analysis was performed using R4.2.2 software; the remaining analyses were performed using IBM SPSS statistical software v26.0 and GraphPad Prism 8.0. The test level was set at α = 0.05.

The data that support the findings of this study are available from the corresponding author upon reasonable request.

## RESULTS

### ApolJ-Glyc was associated with the diagnosis of AMI

All enrolled patients were included in Control group (n=88), AMI group (n=75). Table 1 summarizes the baseline data characteristics of the two groups of patients.

**Table 1.**
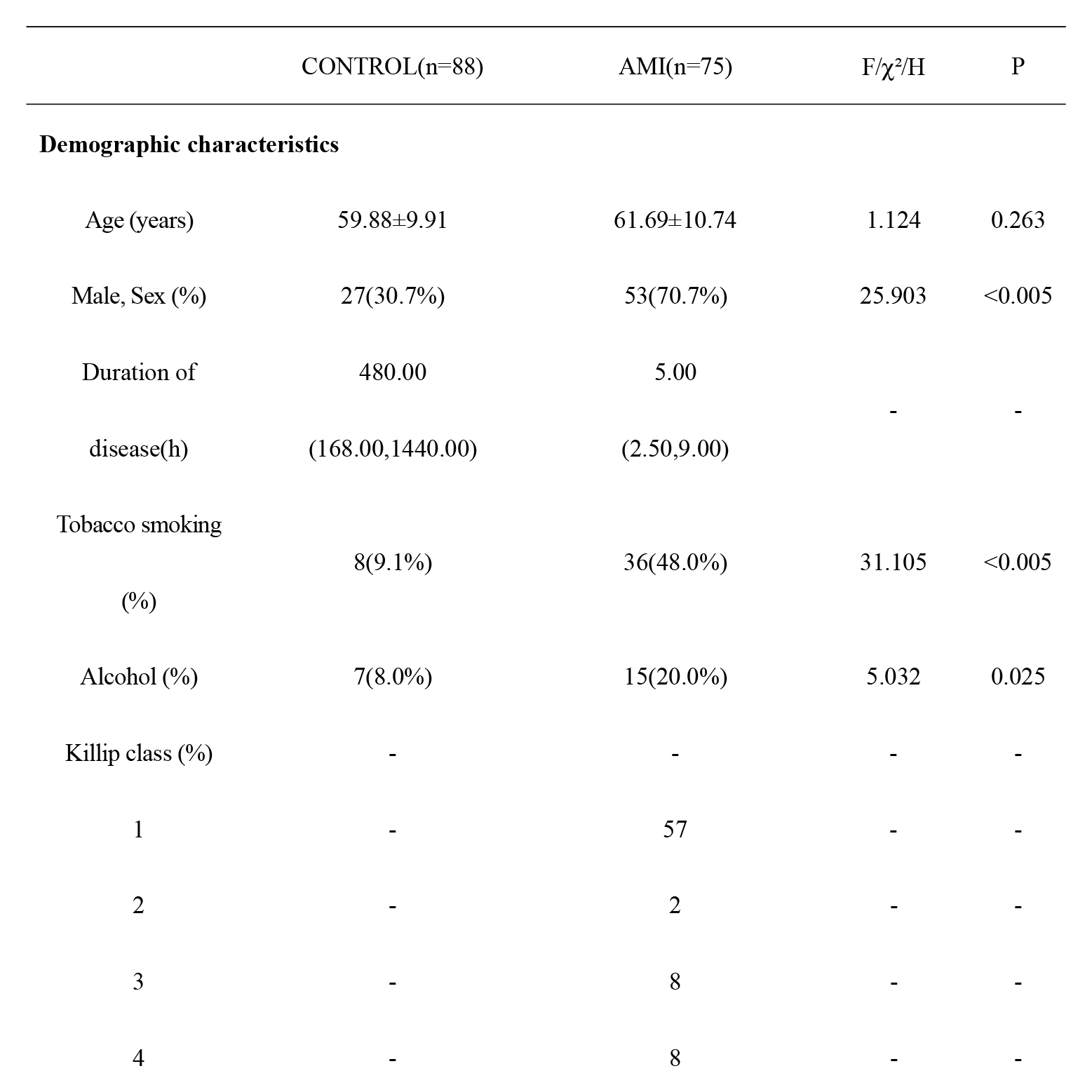
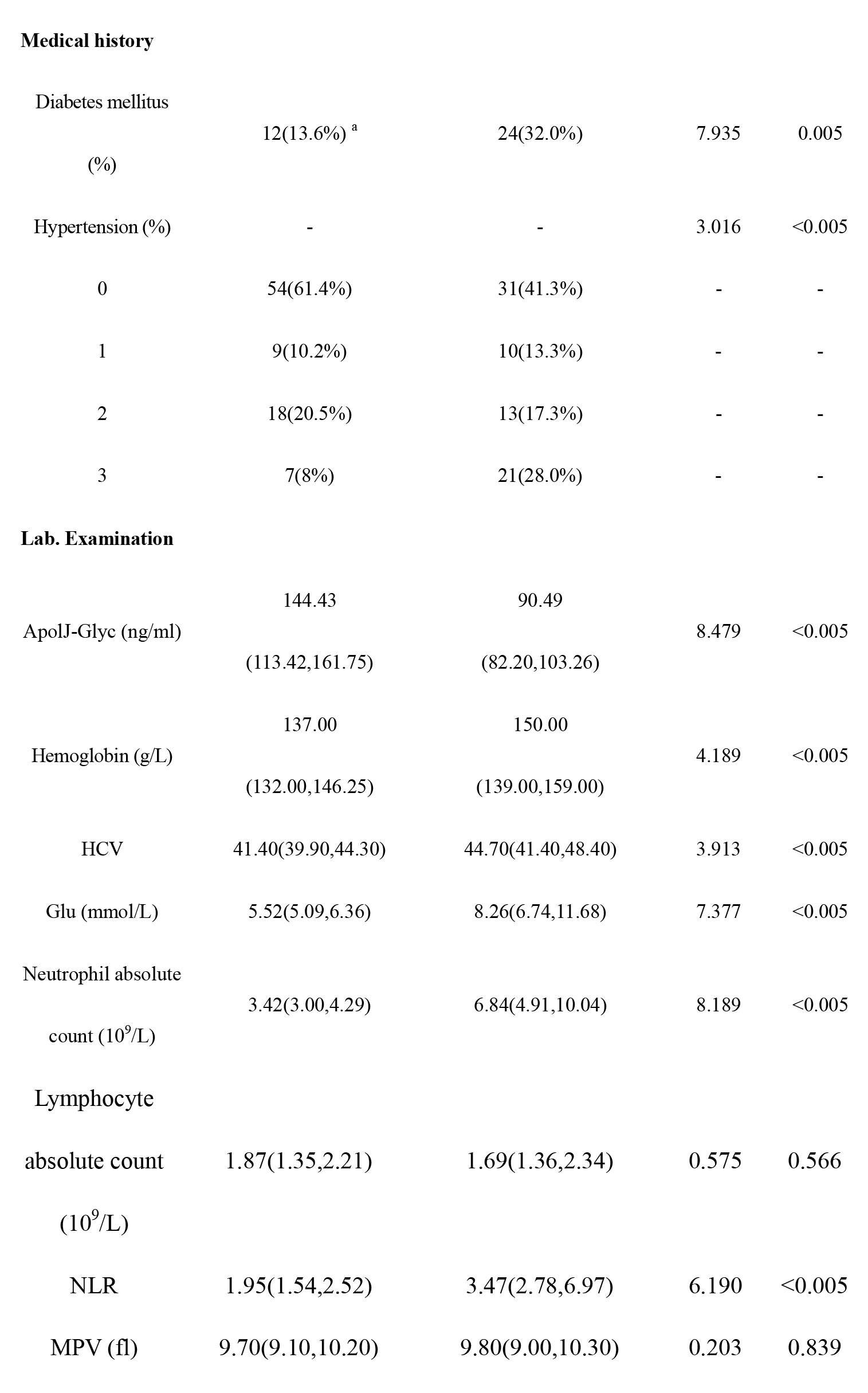
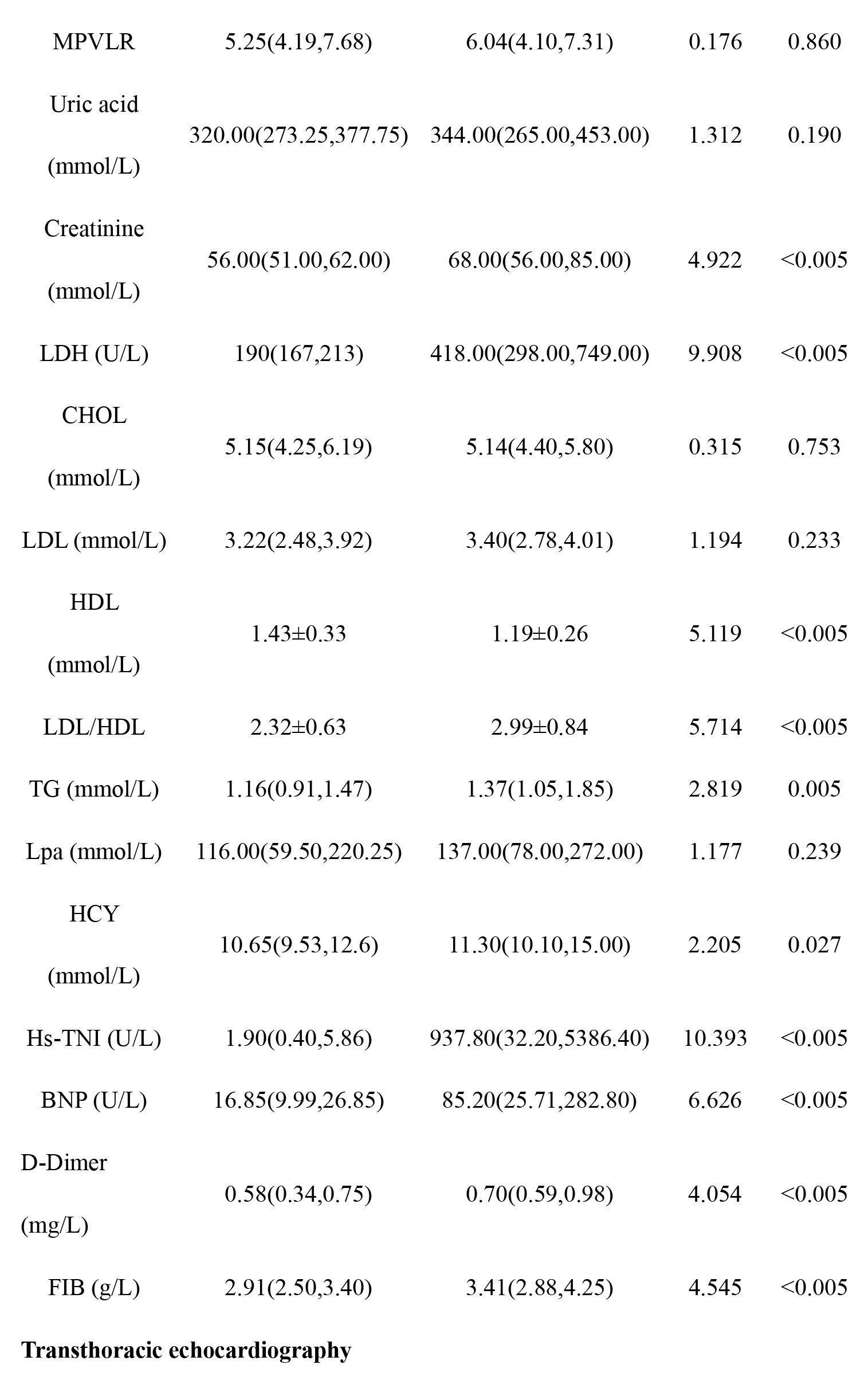
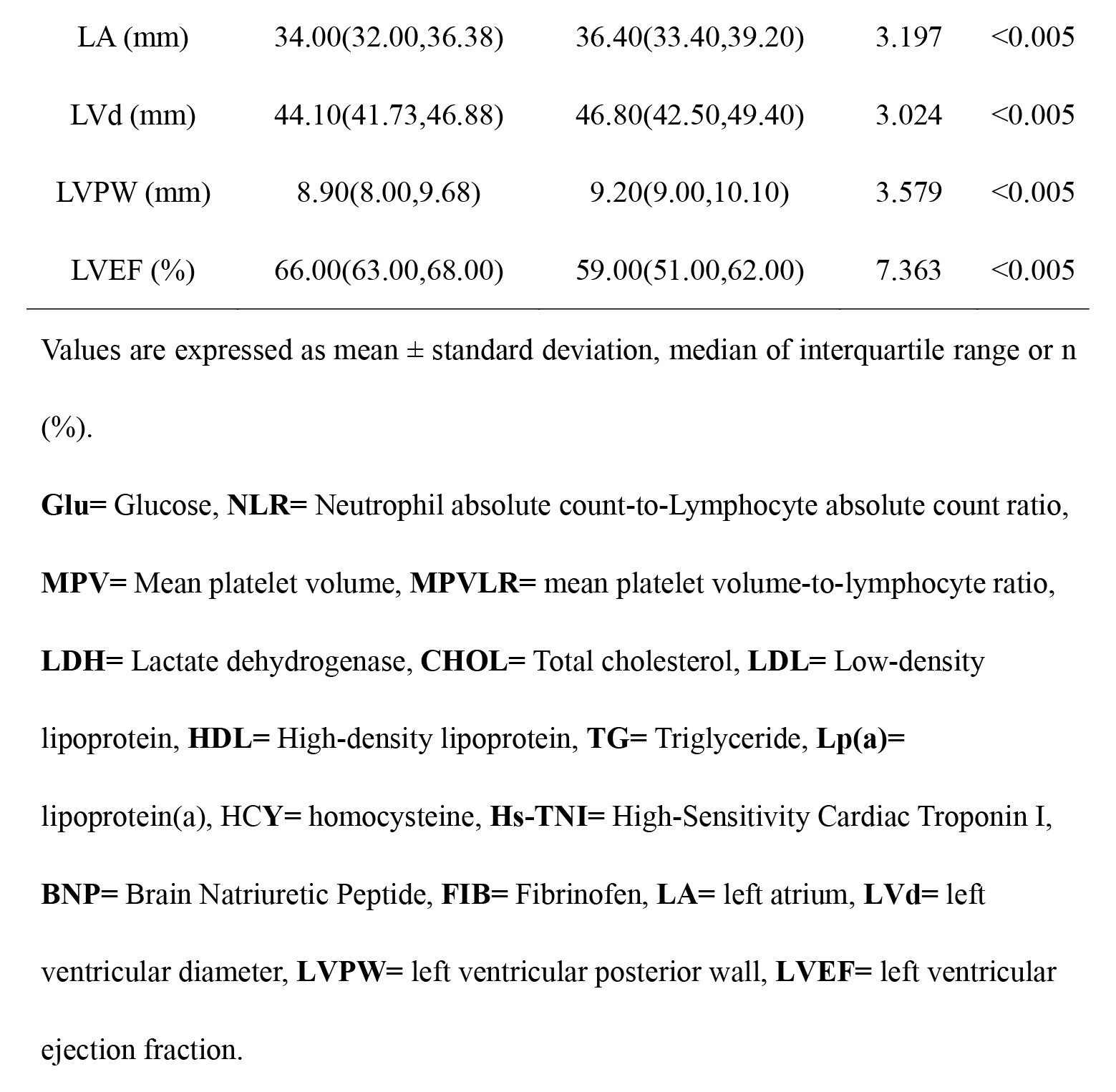
Baseline characteristics of all subjects.

The results showed that the AMI group and the control group were statistically different in terms of gender, smoking, history of alcohol consumption, diabetes, hypertension, apolJ-Glyc, HB, HCV, glu, absolute neutrophil count, NLR, creatinine, LDH, HDL, LDL/HDL, TG, HCY, hsTNI, BNP, D-D, FIB, LA, LVd LVPW, and LVEF were all statistically significant differences when compared in these variables. (P<0.05)

Based on the results of the univariate analysis of variance, a R language-Lasso model was developed for 25 statistically significant variables (p < 0.05), and the LASSO screening showed that the number of retained variables was 12, namely gender, smoking, absolute neutrophil count, NLR, hsTnI, FIB, glu, LDH, LDL/HDL, ApolJ-Glyc, and LVPW, LVEF. (Figure 1)

**Figure 1.**
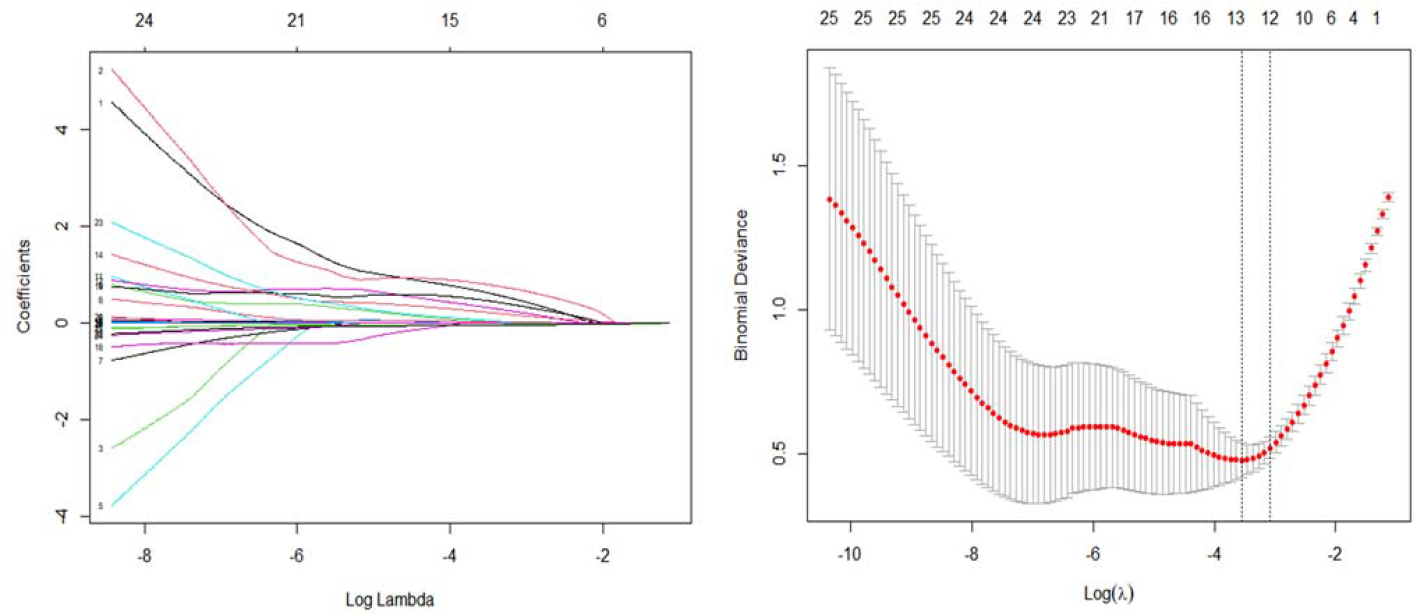
Lasso model development and variable screening. LASSO screening showed that lambda.min=12, namely gender, smoking, absolute neutrophil count, NLR, hsTnI, FIB3, glu, LDH, LDL/HDL, ApolJ-Glyc, and LVPW, LVEF.

Then, these 12 variables were included in a multifactorial logistic regression model (mode: forward method) and the variables retained in the final model were smoking, hsTnI, LDH, APOLJ-Glyc (p < 0.05)

The results revealed that smoking (OR 10.976,95% CI 1.105∼108.985, P=0.041), hsTnI(OR 1.061,95% CI 1.001∼1.125, P=0.047), LDH(OR 1.030,95% CI 1.011∼1.050, P=0.002), and APOLJ-Glyc(OR 0.953,95% CI 0.912∼0.997, P=0.038) were independent risk factors for acute myocardial infarction, elevated serum ApolJ-Glyc levels were protective factors for acute myocardial infarction, and smoking, elevated serum LDH and hsTnI were risk factors for acute myocardial infarction (P < 0.05) (Table 2).

**Table 2.**
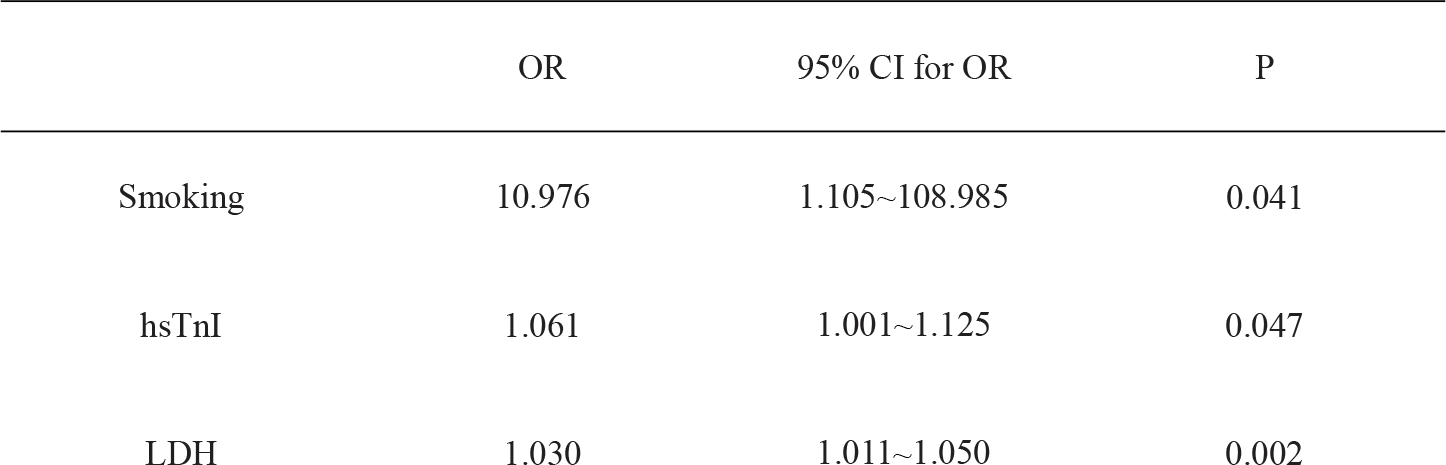
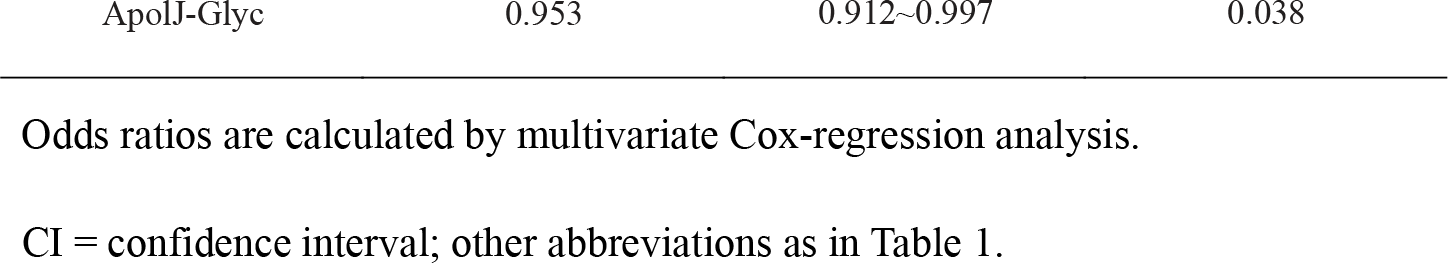
Risk factors for acute myocardial infarction.

Besides, ROC analysis was performed on this regression model, and the results showed that : The model showed a very high discriminatory value for acute myocardial infarction, with an area under the curve (AUC) of 0.995, a sensitivity of 99% and a specificity of 97% (P< 0.0001) (Table 3).

**Table 3.** The ROC curves for the multivariate analysis

|  | AUC | Sensitivity (%) | Specificity (%) | Youden index | Cut-V | p |
| --- | --- | --- | --- | --- | --- | --- |
| Smoking | 0.695 | 48 | 91 | 0.573 | - | 0.000 |
| LDH | 0.951 | 89 | 92 | 0.813 | 246.500 | 0.000 |
| ApoJ-Glyc | 0.886 | 89 | 72 | 0.606 | 99.240 | 0.000 |
| hs-TnI | 0.973 | 91 | 98 | 0.884 | 15.800 | 0.000 |
| multiple<br>actor | 0.995 | 99 | 97 | 0.964 | - | 0.000 |

AUC= Area Under Roc, other abbreviations as in Table 1.

Meanwhile, compared to controls at admission, serum APOLJ-Glyc levels (t=0) were 37% lower in patients with AMI (median: AMI (n= 75): 90.49 vs Control (n = 88): 144.43ng/ml; p< 0.0001). ROC analysis showed that the measurement of ApoJ-Glyc levels was significant for the identification of acute myocardial infarction, with an area under the curve (AUC) of 0.886, a cut-value of 99.24ng/ml, a sensitivity of 89% and a specificity of 72% (p< 0.0001) (Figure 2).

**Figure 2.**
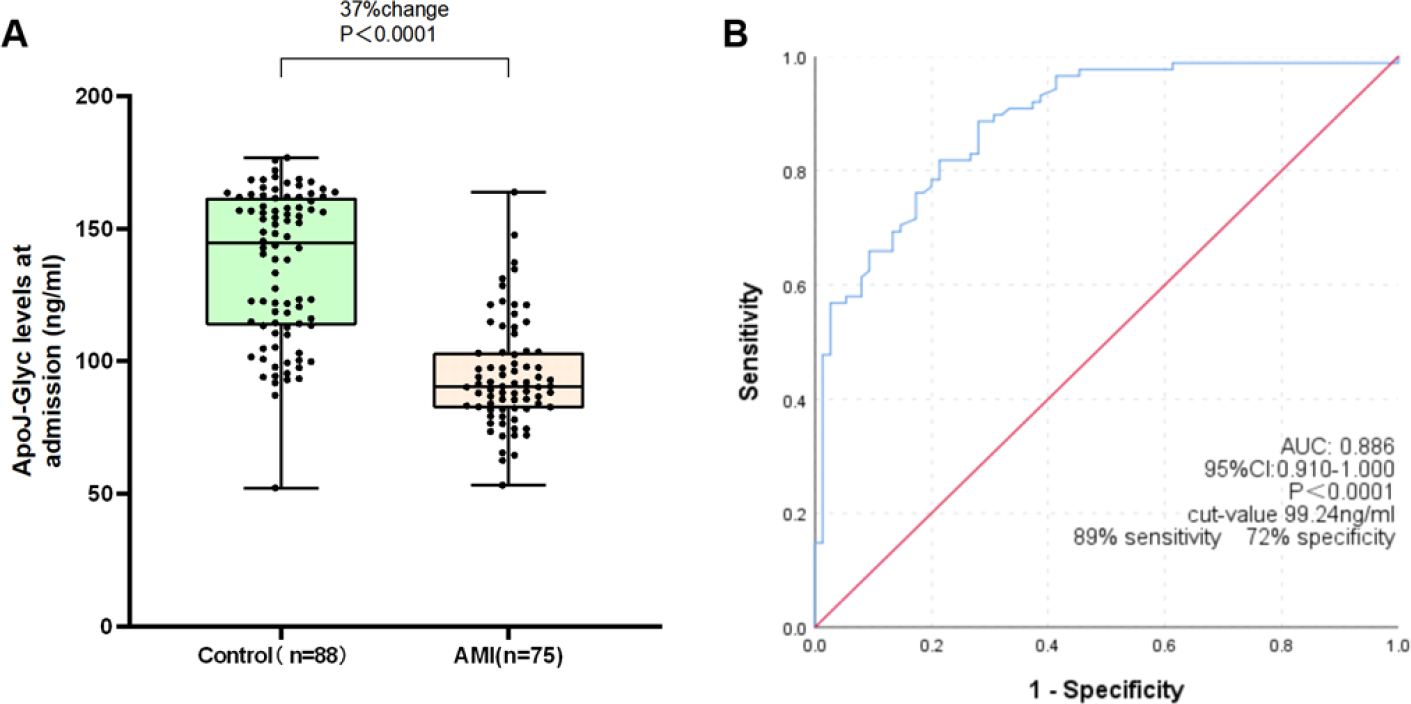
Glycosylated apolipoprotein J diagnostic value. (A) Box plot showing glycosylated apolipoprotein J levels in acute myocardial infarction patients (n= 75) and in healthy subjects (n= 88). (B) Glycosylated apolipoprotein J levels have a diagnostic value for acute myocardial infarction, as shown by a receiver operating characteristic curve with an area under the curve of 0.886. (P< 0.0001) and a cut-off value of 99.24ng/ml with 89% of sensitivity and 72% of specificity.

Of the 75 patients collected with acute infarction who underwent coronary stenting, they were divided into NSTEMI and STEMI groups according to ECG criteria. There were statistical analyses of four variables, smoking, hsTnI, LDH and APOLJ-Glyc, between the NSTEMI group, the STEMI group and the CONTROL group.

The results showed that there was no statistically significant comparison between the NSTEMI and STEMI groups in the four variables of smoking, apolJ-Glyc, hsTNI and LDH, but both groups were statistically different from the control group (P < 0.001) (Table 4).

**Table 4.** Comparison between groups based on independent risk factors.

| | CONTROL(n=88) | NSTEMI(n=37) | STEMI(n=38) | F/$\chi^2$/H | p |
| --- | --- | --- | --- | --- | --- |
| Smoking | 8 (9.1%)<sup>a</sup> | 20(54.1%)<sup>b</sup> | 16(42.1%)<sup>b</sup> | 32.463 | 0.000 |
| (%) |  |  |  |  |  |
| ApoJ-Glyc | 144.43 | 90.63 | 90.24 | 71.968 | 0.000 |
| (ng/ml) | (113.42,161.75)<sup>a</sup> | (84.73,103.58)<sup>b</sup> | (78.42,,103.10)<sup>b</sup> |  |  |
| LDH(U/L) | 190 (167,213)<sup>a</sup> | 376(268,558)<sup>b</sup> | 560(355,1086)<sup>b</sup> | 101.669 | 0.000 |
| hsTNI(U/L) | 1.9 (0.40,5.86)<sup>a</sup> | 3066.70(82.55,14519.05)<sup>b</sup> | 463.50(27.90,2350.45)<sup>b</sup> | 108.896 | 0.000 |

Note: Values are expressed as mean ± standard deviation, median of interquartile

Patients with early myocardial infarction are defined as, at the onset of symptoms until the first blood sample taken upon admission, the conventional hs-TnI level in patients with acute myocardial infarction was negative, and beyond that, the hsTnI was higher than the reference value at the 99th percentile. The early MI group contained 22 patients from the STEMI group and NSTEMI group, the rest was late MI group(n=53).To investigate the discriminatory value of ApolJ-Glyc in different periods for acute myocardial infarction, according to the criteria, all enrolled patients were included in the Early-MI group (n=22) and Late-MI group (n=53), stratified analysis with Control group (n=88) respectively.

### Glycosylated apolipoprotein J in Early Myocardial Infarction

The 12 variables retained from the LASSO screening, namely gender, smoking, absolute neutrophil count, NLR, hsTnI, FIB, glu, LDH, LDL/HDL, ApolJ-Glyc, LVPW, LVEF, were included in a multifactorial logistic regression model (model: forward approach), and the variables retained in the final model were smoking, LDH, and APOLJ-Glyc (p < 0.05).The results showed that smoking(OR 15.694,95% CI 1.790∼137.581), LDH(OR 1.030,95% CI 1.012∼1.049), and APOLJ-Glyc(OR 0.958,95% CI 0.912∼0.997) were independent risk factors for early myocardial infarction, elevated serum ApolJ-Glyc levels were protective factors for acute myocardial infarction, and smoking and elevated serum LDH were risk factors for early myocardial infarction (P < 0.05) (Table 5).

**Table 5.** Risk factors for early myocardial infarction

|  | OR | 95% CI for OR | P |
| --- | --- | --- | --- |
| Smoking | 15.694 | 1.790~137.581 | 0.013 |
| LDH | 1.030 | 1.012~1.049 | 0.001 |
| ApoJ-Glyc | 0.958 | 0.912~0.997 | 0.036 |

Abbreviations as in Table 1.

As well, ROC analysis was performed on this regression model, and the results showed that : The model showed a high discriminatory value for early myocardial infarction, with an area under the curve (AUC) of 0.980, a sensitivity of 96% and a specificity of 98% (P< 0.0001) (Table 6).

**Table 6.** The ROC curves for the multivariate analysis.

|  | AUC | Sensitivity (%) | Specificity (%) | Youden index | Cut-V | p |
| --- | --- | --- | --- | --- | --- | --- |
| Smoking | 0.773 | 64 | 91 | 0.50 | - | 0.000 |
| LDH | 0.954 | 91 | 92 | 0.829 | 246.5 | 0.000 |
| ApoJ-Glyc | 0.871 | 76 | 82 | 0.579 | 113.33 | 0.000 |
| multiple<br>actor | 0.980 | 96 | 98 | 0.932 | - | 0.000 |

Abbreviations as in Table 1 and Table 3.

Meanwhile, compared to control group, in patients with early-AMI, ApoJ-Glyc levels were 36% lower on admission before myocardial injury markers (hsTnI) were elevated (median: E-AMI (n= 22): 92.98 vs Control (n = 88): 144.43ng/ml; P< 0.0001). ROC analysis showed that ApoJ-Glyc levels were also important in the early identification of acute myocardial infarction, with an area under the curve (AUC) of 0.871 (P< 0.0001), a critical value of 113.33ng/ml, a sensitivity of 76% and a specificity of 82% (Figure 3).

**Figure 3.**
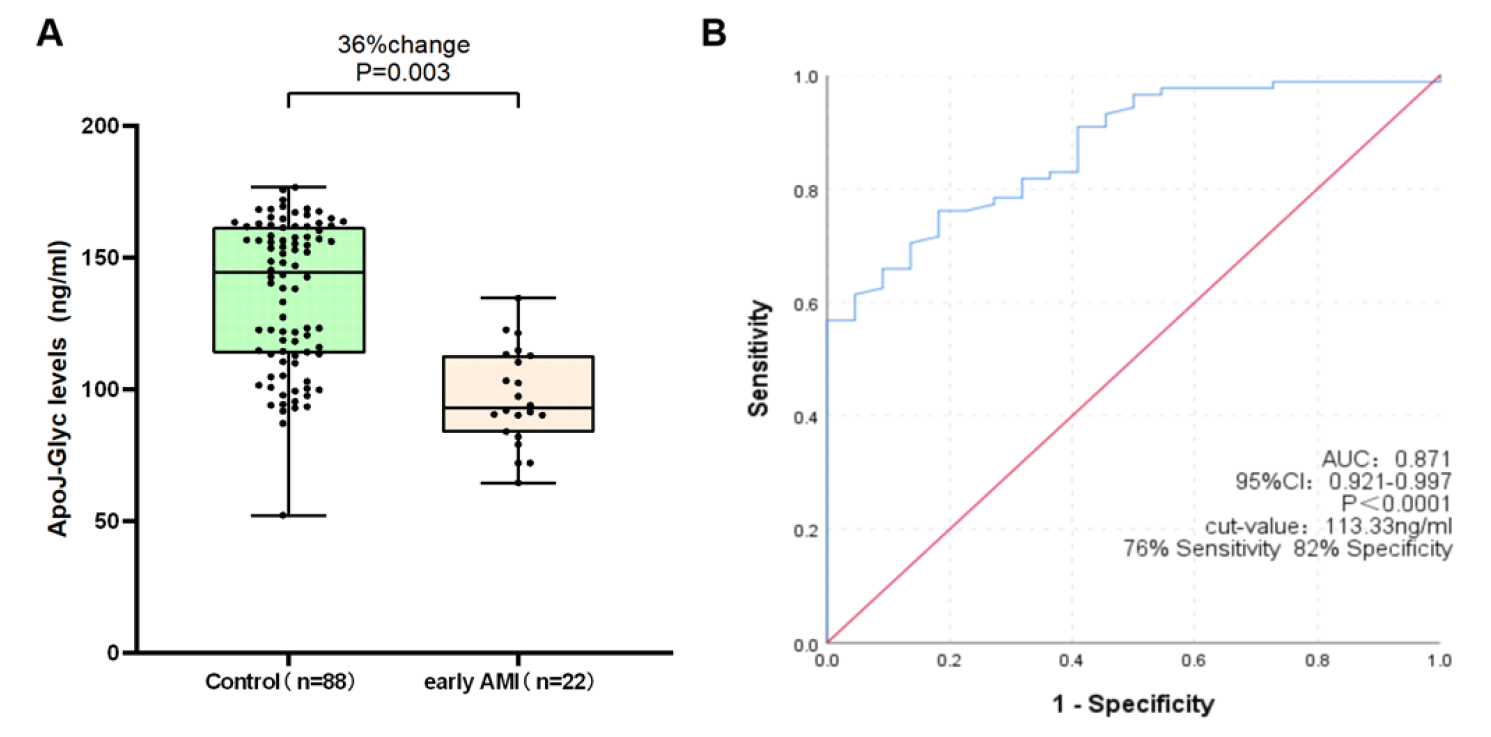
Glycosylated apolipoprotein J diagnostic value. (A) Box plot showing glycosylated apolipoprotein J levels in early acute myocardial infarction patients (n= 22) and in healthy subjects (n= 75). (B) Glycosylated apolipoprotein J levels have a diagnostic value for acute myocardial infarction, as shown by a receiver operating characteristic curve with an area under the curve of 0.871. (P< 0.0001) and a cut-off value of 113.33ng/ml with 76% of sensitivity and 82% of specificity.

### Glycosylated apolipoprotein J in Late Myocardial Infarction

The 12 variables retained from the LASSO screening, namely gender(males), smoking, absolute neutrophil count, NLR, hsTnI, FIB, glu, LDH, LDL/HDL, ApolJ-Glyc, LVPW, LVEF, were included in a multifactorial logistic regression model (model: forward approach), and the variables retained in the final model were males, NLR, FIB, Glu, ApolJ-Glyc, LVPW, LVEF. (p < 0.05).The results showed that males(OR 122.488,95%CI 1.314∼11417.973),NLR(OR 3.020,95% CI 1.106∼8.245), FIB(OR 25.887,95% CI 1.895∼353.659),Glu(OR 4.858,95% CI 1.404∼16.813), ApolJ-Glyc(OR 0.909,95% CI 0.854∼0.968), LVPW(OR 15.957,95% CI 1.517∼167.817), LVEF(OR 0.598,95% CI 0.401∼0.892) were independent risk factors for late myocardial infarction, elevated serum ApolJ-Glyc levels and LVEF were protective factors for late myocardial infarction, and males, elevated serum NLR, FIB, Glu levels and LVPW were risk factors for late myocardial infarction (P < 0.05) (Table 7).

**Table 7.** Risk factors for late myocardial infarction.

|  | OR | 95% CI for OR | P |
| --- | --- | --- | --- |
| Males | 122.488 | 1.314~11417.973 | 0.038 |
| NLR | 3.020 | 1.106~8.245 | 0.031 |
| FIB3 | 25.887 | 1.895~353.659 | 0.015 |
| Glu | 4.858 | 1.404~16.813 | 0.013 |
| ApolJ-Glyc | 0.909 | 0.854~0.968 | 0.003 |
| LVPW | 15.957 | 1.517~167.817 | 0.021 |
| LVEF | 0.598 | 0.401~0.892 | 0.012 |

Abbreviations as in Table 1 and Table 2.

As well, ROC analysis was performed on this regression model, and the results showed that: The model showed a very high discriminatory value for late myocardial infarction, with an area under the curve (AUC) of 0.998, a sensitivity of 99% and a specificity of 91% (P< 0.0001) (Table 8).

**Table 8.** The ROC curves for the multivariate analysis.

|  | AUC | Sensitivity (%) | Specificity (%) | Youden index | Cut-V | p |
| --- | --- | --- | --- | --- | --- | --- |
| Males | 0.677 | 66 | 69 | 0.500 | - | 0.000 |
| NLR | 0.836 | 74 | 83 | 0.566 | 2.75 | 0.000 |
| FIB3 | 0.757 | 45 | 96 | 0.408 | 4.02 | 0.000 |
| Glu | 0.835 | 76 | 84 | 0.596 | 6.74 | 0.000 |
| ApoJ-Glyc | 0.892 | 89 | 77 | 0.660 | 99.24 | 0.000 |
| LVPW | 0.676 | 79 | 55 | 0.326 | 8.95 | 0.000 |
| LVEF | 0.881 | 92 | 74 | 0.656 | 60.5 | 0.000 |
| multiple<br>actor | 0.998 | 99 | 91 | 0.909 | - | 0.000 |

AUC=Area Under Roc, other abbreviations as in Table 1.

Compared to the control group, patients with L-AMI (the hsTnI was higher than the reference at the 99th percentile) had a 39% reduction in ApoJ-Glyc levels at admission (median: L-AMI (n= 53): 88.14 vs Control (n = 88): ROC analysis showed that the measurement of ApoJ-Glyc levels was also important in the identification of acute myocardial infarction after a significant increase in myocardial injury markers, with a greater area under the curve (AUC) of 0.892 (P< 0.0001), a critical value of 99.24ng/ml, a sensitivity of 89% and a specificity of 77% (Figure 4).

**Figure 4.**
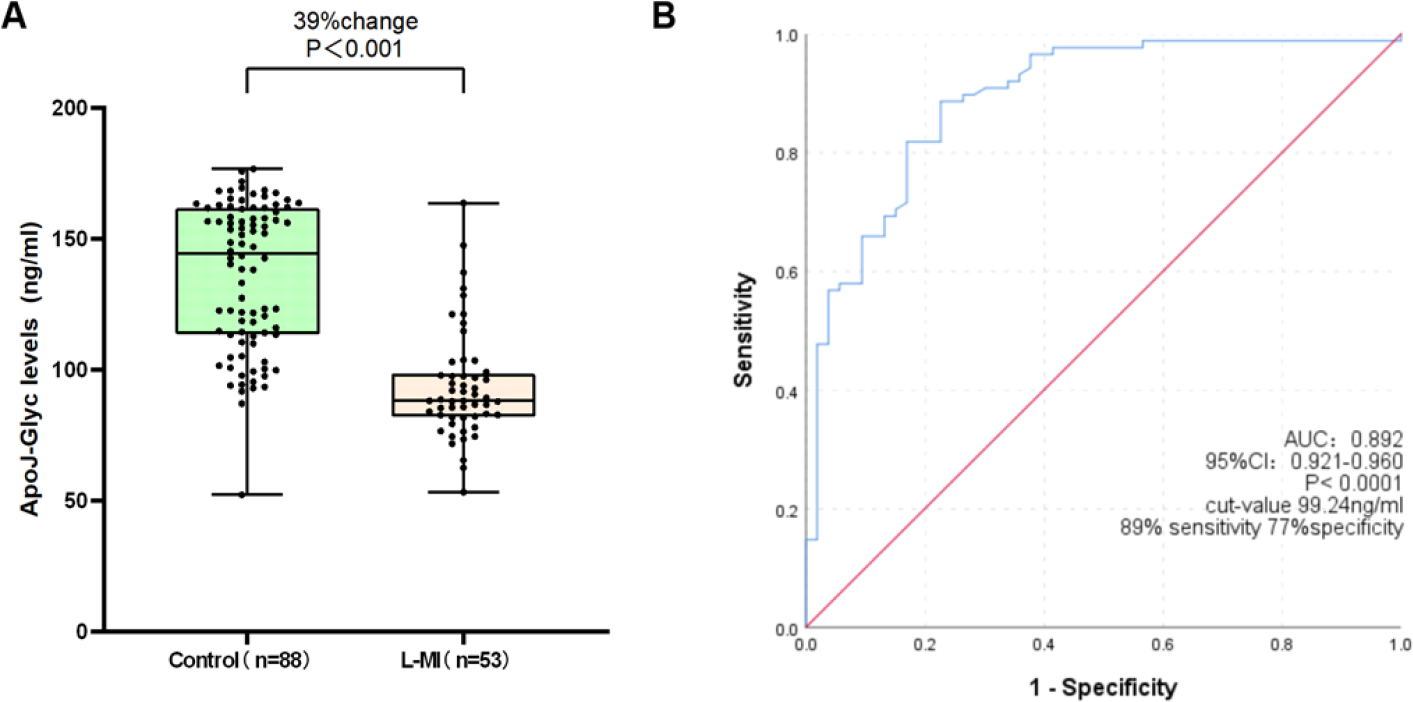
Glycosylated apolipoprotein J diagnostic value. (A) Box plot showing glycosylated apolipoprotein J levels in late myocardial infarction patients (n= 53) and in healthy subjects (n= 75). (B) Glycosylated apolipoprotein J levels have a diagnostic value for late myocardial infarction, as shown by a receiver operating characteristic curve with an area under the curve of 0.892. (P< 0.0001) and a cut-off value of 99.24ng/ml with 89% of sensitivity and 77% of specificity.

### ApolJ-Glyc Was Associated With the Severity of AMI

Statistically significant variables in Table1 and indicators to evaluate the severity of MI were subjected to spearman correlation analysis with ApolJ-Glyc and screened for statistically significant indicators (p<0.05); The results showed an positive correlation between ApolJ-Glyc and HDL and TIMI flow grade after PCI, as well as a negative correlation between ApolJ-Glyc and creatinine, KILLIP grade, and Gensini scores (Table 9).

**Table 9.** Spearman correlation between indicator and ApolJ-Glyc in the AMI group.

| Indicators | rs | p |
| --- | --- | --- |
| Sex (male) | 0.080 | 0.492 |
| Age (year) | -0.097 | 0.408 |
| Smoking (%) | 0.022 | 0.850 |
| Alcohol (%) | 0.138 | 0.238 |
| Diabetes (%) | 0.080 | 0.496 |
| hsTnI | -0.103 | 0.380 |
| Killip grade | -0.256 | 0.027 |
| GLU | -0.102 | 0.383 |
| NLR | -0.015 | 0.901 |
| FIB3 | -0.171 | 0.143 |
| creatinine | -0.234 | 0.044 |
| HDL | 0.297 | 0.010 |
| LDL/HDL | -0.043 | 0.714 |
| EF (%) | 0.179 | 0.123 |
| Gensini | -0.240 | 0.038 |
| TIMI flow after PCI | 0.355 | 0.002 |

Abbreviations as in Table 1.

Part 1 ApolJ-Glyc Was Associated With the KILLIP.

The KIllIP cardiac function classification is an important index for evaluating cardiac impairment in patients with acute myocardial infarction. Analysis of the differences in ApolJ-Glyc levels in the AMI population with different KillIP classifications showed statistically significant results (P = 0.021) and were compared between groups, with no statistical significance between groups with KillIP classifications of 1, 2 and 3 (see supplementary material), analyses performed after combining the groups showed that serum ApoJ-Glyc levels (t = 0) were lower in patients with AMI combined with cardiogenic shock on admission compared with patients with myocardial infarction without cardiogenic shock (P = 0.003) (Figure 5A).

Part 2 ApolJ-Glyc Was Associated With the TIMI flow.

**Figure 5.**
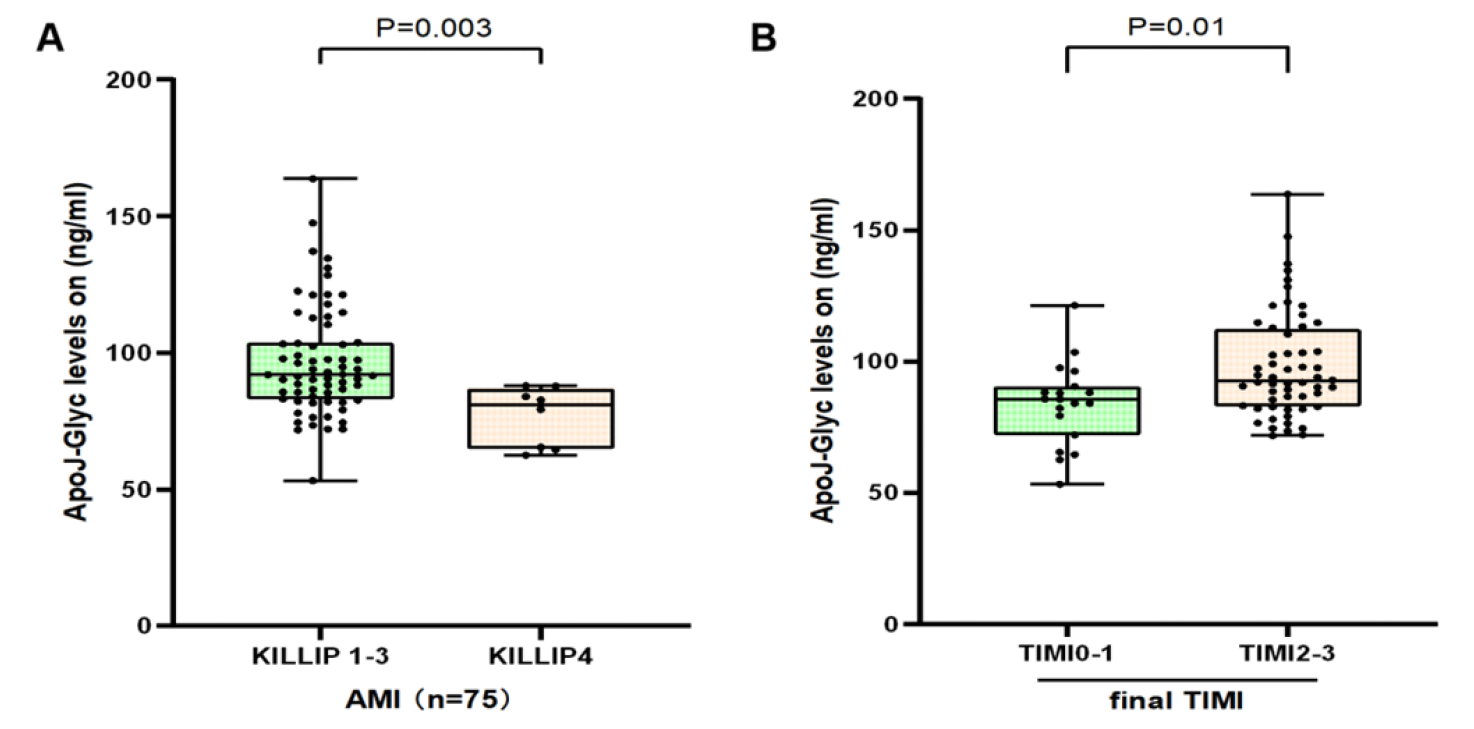
Glycosylated apolipoprotein J assess the severity of AMI. (A) Box plot depicting the notable differences in glycosylated apolipoprotein J levels at admission based on the killip cardiac function classification. (B) Box plot depicting the notable differences in glycosylated apolipoprotein J levels at admission based on the final TIMI flow grade.

TIMI flow grading evaluates distal blood flow in diseased vessels in patients with myocardial infarction. The current study has shown that a TIMI flow grade of 0 or 1 after PCI may mean a worse prognosis compared to patients with a TIMI flow grade of 2 or 3 after PCI29. Statistical analysis showed that the group with a final TIMI flow≤1 showed lower serum ApoJ-Glyc levels (t=0)compared to the group with a final TIMI flow≥2 (P=0.010) (Figure 5B).

Part3 ApolJ-Glyc Was Associated With the Gensini score.

The Gensini score evaluates the degree of coronary artery lesion stenosis in patients and linear regression analysis shows that Gensini score is independent of ApolJ-Glyc (Durbin-Watson: 1.492).Moreover serum ApoJ-Glyc levels (t=0)has predictive value for thrombotic load in AMI patients: the lower the ApolJ-Glyc level, the heavier the thrombotic load in AMI patients is likely to be (β=-0.442, P<0.05).

### ApolJ-Glyc Was Associated With the prognosis of AMI

Serum ApoJ-Glyc levels were quantified in serum samples 72 hours after admission in 69 patients with AMI (blood samples could be collected 72 hours after admission). Serum ApoJ-Glyc levels recovered compared to the levels at admission. (Median: t=72h: 105.76 vs. t=0: 90.49ng/ml; p< 0.05)(Figure 6A). When the changes in ApoJ-Glyc levels were analysed separately, two different groups of patients were defined, those with stable or even increasing (recovering) ApoJ-Glyc levels and those with progressively decreasing ApoJ-Glyc levels (p< 0.05)(Figure 6B). Patients with progressively decreasing serum ApoJ-Glyc levels had a higher probability of MACCE 6 months after discharge compared to patients with stable or even increasing (recovering) serum ApoJ-Glyc levels(p< 0.05)(Figure 6C).Survival curves differed between the group with stable or even increasing (recovering) ApoJ-Glyc levels and the group with progressively decreasing ApoJ-Glyc levels (Log Rank P < 0.001), and patients in the group with progressively decreasing ApoJ-Glyc levels did show more MACCE (p< 0.05)(Figure 6D).

**Figure 6.**
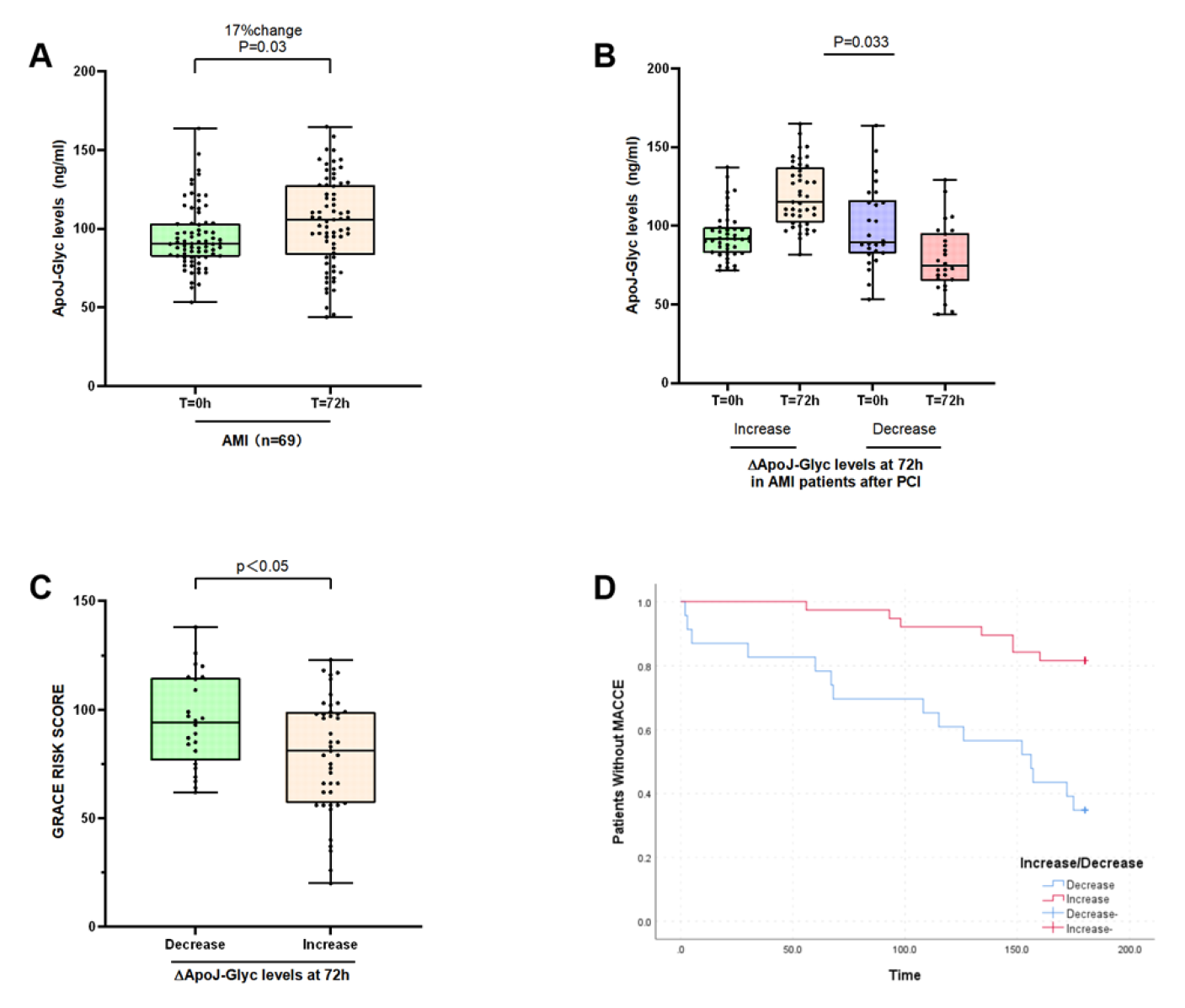
Glycosylated apolipoprotein J assess the prognosis of AMI. (A) Box plot depicting Serum ApoJ-Glyc levels recovered compared to the levels at admission. (B) When the changes in ApoJ-Glyc levels were analysed separately, patients with stable or even increasing (recovering) ApoJ-Glyc levels and those with progressively decreasing ApoJ-Glyc levels. (C) Correlation between GRACE risk score and decreased glycosylated apolipoprotein J levels 3 days after PCI in patients with acute myocardial infarction. (D) Cumulative Kaplan-Meier estimates of the freedom from MACCE.

## DISCUSSION

Acute myocardial infarction (AMI) caused by chest pain is the most severe form of coronary heart disease, which contributes to the leading cause of cardiovascular death and disability and is one of the most common causes of admission worldwide^30^. Meanwhile, increasing evidence indicated that the presence of obstructive coronary artery disease is the strongest predictor of future adverse cardiovascular outcomes^30–32^.

Patients with acute chest pain show obvious heterogeneity. On the other hand, it is important to distinguish myocardial infarction from non-ischemic chest pain^33^.Despite slightly increased levels of coronary revascularization and preventive therapy in patients with acute MI, recommendations based on high sensitivity troponin and universal definition did not improve outcomes and future cardiovascular events did not decrease^34^.Therefore, for patients with acute myocardial infarction, there is an urgent need to encourage clinicians to consider the potential pathological mechanisms of myocardial injury and to find new markers of myocardial injury, rather than just empirically using "troponin" as the gold standard. This benefits disease detection and risk stratification during the early phases of hospitalization, as well as the ability to anticipate the probability of unfavorable outcomes after discharge, all of which are critical for optimizing treatment and reducing ongoing medical costs.

In recent years, it has been found that the different glycosylation states of proteins through processing and modification depend on tissue and disease-specific pathophysiological states^35–40^. The assessment of protein concentrations in various glycosylated forms has emerged as a new technique for diagnosing various disorders. Apolipoprotein J is a molecular chaperone with antioxidant and cytoprotective properties^34^. Studies on apoE^-^ mice and monkeys using apoJ peptide fragments support the potential anti-atherosclerotic effects of clusterin^41^. Notably, recent animal studies suggest that ApoJ-Glyc may be an early and highly sensitive biomarker of acute myocardial infarction with varying degrees of recovery on the third day after myocardial infarction^27^. Hence, we describe the value of ApoJ-Glyc at the human circulatory level in the diagnosis and prognosis of AMI.

### Glycosylated apolipoprotein J: an early alarm bell for an acute myocardial infarction

In this paper, we show that serum levels of ApoJ-Glyc are not only associated with the presence of AMI, but that ApoJ-Glyc is an independent risk factor for AMI. Interestingly, serum ApoJ-Glyc levels did not show significant differences between STEMI and NSTEMI populations, and ApoJ-Glyc levels alone did not distinguish between STEMI and NSTEMI myocardial infarction types, which had not been found in previous studies^42^. Meanwhile, we show that the measurement of serum ApoJ-Glyc levels helped in the early identification of patients with acute myocardial infarction; before the detection of elevated ultrasensitive troponin (between 1.38h and 4h of onset), a significant decrease in serum ApoJ-Glyc levels was detected in AMI populations compared to normal subjects. Similarly, some clinical studies for STEMI patients have also demonstrated that the reduction in ApoJ-Glyc secretion occurs prior to the elevation of ultrasensitive troponin, presumably as evidence of irreversible cellular damage^43^,which is consistent with some of the results of current experiment. In addition, the serum ApoJ-Glyc levels were also important in the identification of acute myocardial infarction after a significant increase in myocardial injury markers. Consequently, all these results suggest that glycosylated apolipoprotein J plays an important warning role in acute myocardial infarction.

### Glycosylated apolipoprotein J after acute myocardial infarction

After early diagnosis and treatment of serious diseases, timely assessment of residual risk of serious illness, active intervention and follow-up all remain crucial. An unanticipated finding is that there is a correlation between serum ApoJ-Glyc levels in patients with acute myocardial infarction at emergency admission and indicators that evaluate the severity of myocardial infarction (such as KILLIP classification, Gensini score, TIMI blood flow classification). The lower the serum ApoJ-Glyc level, the more severe the patient may be. Unexpectedly, this study showed a clear correlation between serum glycosylated apolipoprotein J levels and Gensini scores. It can be seen that serum glycosylated apolipoprotein J levels have some predictive value for the severity of coronary lesions in patients with myocardial infarction. However, due to the limited sample size of this study, further studies are needed to determine whether serum glycosylated apolipoprotein J levels are predictive of the severity of coronary artery disease in patients with other types of ACS^44^. Obviously, by combining ApoJ-Glyc levels at admission, a joint assessment of the patient’s condition and in-hospital risk of adverse events could conceivably help to implement more individualized treatment and even improve risk stratification.

More meaningfully, changes in serum ApoJ-Glyc levels were still detectable after acute onset. Firstly, the assessment of trends in serum ApoJ-Glyc levels 72h after admission to hospital indicated that changes in ApoJ-Glyc levels have a potential role in the progression of the disease. Furthermore, a sustained decrease in serum ApoJ-Glyc levels may be directly associated with a worse prognosis. Moreover, we followed AMI patients for 6 months using the presence of MACCE within 6 months as an endpoint and the observed gradual decrease in ApoJ-Glyc levels after 72h of admission did result in more MACCE, consistent with the predictions of higher GRACE risk score. Previous clinical studies on selected STEMI patients have shown that a gradual decrease in ApoJ-Glyc levels after admission may be correlated with higher GRACE scores^43^. Even though we did not replicate the previous studies, it is clear that our results have contributed to a profound and novel understanding of the function of ApoJ-Glyc: 1) a progressive decrease in ApoJ-Glyc levels is linked to worse prognosis and poorer outcomes, 2) the links between changes in ApoJ-Glyc levels and prognosis also highlight the potential role of ApoJ-Glyc at circulating levels at different stages after the onset of acute myocardial infarction.

## CONCLUSION

This study showed a correlation between serum ApoJ-Glyc and AMI:

1. Serum ApoJ-Glyc is valuable for both early diagnosis and diagnosis of AMI patients.
2. Serum ApoJ-Glyc did not show significant variability between STEMI patients and NSTEMI patients.
3. Serum ApoJ-Glyc levels were significantly associated with the presence and severity of AMI.
4. Changes in serum ApoJ-Glyc levels were significantly associated with ischaemic risk assessment and MACCE after AMI.

### Limitation

1. Insufficient sample size and lack of evaluation of its performance in large prospective clinical validation trials and adequate assessment of its potential added value in the context of hs-Tn,
2. Clinical value of ApoJ-Glyc in the context of possible other possible mechanisms of cardiac ischaemia, such as non-obstructive coronary artery disease and other non-cardiac factors affecting coronary blood flow dysfunction.

## Nonstandard Abbreviations and Acronyms

ApoJ-Glyc: glycosylated apolipoprotein J

## Data Availability

The data underlying this article will be shared on reasonable request to the corresponding author.

## Acknowledgement

We thank than the following persons for help during the study. YR designed the study. YR, DHB, FKX, JWJ,ZXF performed the experiments. RMM, YR analyzed the data. ZL, GL, WH, JWJ and YR prepared the manuscript. DXN and WAY helped complete ECG. WH helped complete preoperative the transthoracic echocardiographic. All authors have seen and approved the final version of this manuscript.

